# Evaluation of the Roche SARS-CoV-2 Rapid Antibody Test in samples from vaccinated individuals

**DOI:** 10.1101/2021.12.17.21267927

**Authors:** Johannes Hayer, Eva Urlaub

**Affiliations:** Roche Diagnostics GmbH, Mannheim, Germany

**Keywords:** antibodies, neutralizing, COVID-19 vaccines, immunity, immunoassay, nucleocapsid protein, point-of-care systems, SARS-CoV-2, spike protein

## Abstract

**Objective:** The study aimed to establish the performance of the SARS-CoV-2 Rapid Antibody Test (IgG and IgM) and the Elecsys^®^ Anti-SARS-CoV-2 S assay in vaccinated individuals.

**Methods:** A panel of serum samples from Boca Biolistics was utilized to assess antibodies following vaccination, consisting of samples drawn prior to vaccination, after the first dose, or at least 14 days after the second dose of Moderna mRNA-1273 or Pfizer-BioNTech BNT162b2 COVID-19 vaccines. Agreement between the two methods was measured and stratified by test evaluator and assay lot.

**Results:** Agreement between the SARS-CoV-2 Rapid Antibody Test (IgG) and Elecsys Anti-SARS-CoV-2 S assay qualitative measurements at the different assessment points for both mRNA-1273 and BNT162b2 ranged between 97.06% (95% confidence interval [CI] 84.67, 99.93) to 100% (95% CI 82.35, 100). Agreement of the SARS-CoV-2 Rapid Antibody Test (IgG) with the Elecsys Anti-SARS-CoV-2 S assay was not highly influenced by either lot or evaluator. There was a medium-to-strong correlation between the semi-quantitative SARS-CoV-2 Rapid Antibody Test (IgG) result and quantitative Elecsys Anti-SARS-CoV-2 S assay in samples taken after both doses of the vaccines, with higher intensity bands being associated with higher total anti-S antibody titer (mRNA-1273, p=0.0019; BNT162b2, p<0.0001).

**Conclusion:** Semi-quantitative SARS-CoV-2 Rapid Antibody Test (IgG) and quantitative Elecsys Anti-SARS-CoV-2 S assay correlated well, suggesting that the SARS-CoV-2 Rapid Antibody Test (IgG) is helpful in understanding the immune response post-vaccination. The current data support the use of the SARS-CoV-2 Rapid Antibody Test (IgG) in the vaccinated population.

**Importance:** Serologic assays are an essential tool for seroprevalence surveys, for quality control of vaccines, and to determine the response to vaccination. Although a correlate of immunity has not yet been established for COVID-19 vaccines, antibody titers after natural infection and vaccination have been associated with protection from symptomatic SARS-CoV-2 infection. Rapid point-of-care assays can be of use in this context with advantages over centralized testing, such as speed and ease of use. The point-of-care SARS-CoV-2 Rapid Antibody Test (IgG) compared favorably to the Elecsys Anti-SARS-CoV-2 S assay with agreement rates above 97.06%, after one or two doses of Moderna mRNA-1273 or Pfizer-BioNTech BNT162b2. Semi-quantitative SARS-CoV-2 Rapid Antibody Test (IgG) and quantitative Elecsys Anti-SARS-CoV-2 S assay results correlated well, suggesting that SARS-CoV-2 Rapid Antibody Test (IgG) is helpful in understanding the immune response post-vaccination. The current data support the use of the SARS-CoV-2 Rapid Antibody Test (IgG) in the vaccinated population.

## Introduction

Serologic immunoassays are vital in the global management of the COVID-19 pandemic. Measurement of antibodies to SARS-CoV-2 is important for improving disease management, and accurate tests, with validated sensitivity and specificity, are essential for obtaining reliable results when monitoring the pandemic via seroprevalence surveys (1). Serologic assays can provide evidence of recent or past infection (2), and identify those individuals still at risk of infection (3).

Infection with SARS-CoV-2 elicits a strong neutralizing antibody response (4), and antibodies that bind via the spike protein act as neutralizing antibodies (5, 6). Several studies have shown good correlation between anti-S antibodies and functional virus neutralization (7–11). Neutralization assays utilizing live virus require biosafety level 3 containment, which can be labor intensive and low throughput (12–14); therefore anti-S immunoassays, as a surrogate for neutralization, have an important role to play (13, 15). On an individual patient level, anti-S immunoassays can confirm vaccination status or inform upon the need for booster doses (16), which may be helpful for both social and medical reasons.

Moderna mRNA-1273 (spikevax™) and Pfizer-BioNTech BNT162b2 (COMIRNATY^®^) COVID-19 vaccines are mRNA vaccines, approved under the U.S. Food and Drug Administration’s emergency use authorization, each employing a two-dose regimen (17, 18). The immune response involves B cells that produce binding and neutralizing antibodies against the SARS-CoV-2 S protein (19, 20) and both vaccines encode the SARS-CoV-2 spike protein to generate an immune response. Serologic assays are an essential tool in vaccine quality control, being used to determine the response to vaccination and providing insight into the antibody response from the individual patient perspective (21, 22).

The Elecsys^®^ Anti-SARS-CoV-2 S assay (Roche Diagnostics International Ltd, Rotkreuz, Switzerland) is an electrochemiluminescence immunoassay (ECLIA), which has been developed for quantitative *in vitro* detection of antibodies in human serum and plasma (23). The assay detects total antibodies, including immunoglobulin G (IgG), against the receptor binding domain (RBD) of the SARS-CoV-2 spike protein, and is intended for use on the fully automated and high-throughput cobas e analyzers (23). The assay has been shown to detect anti-S antibodies across a variety of different populations (2, 24), and has been utilized as a surrogate of neutralization activity (11, 25).

The SARS-CoV-2 Rapid Antibody Test (Roche Diagnostics International Ltd, Rotkreuz, Switzerland) is CE-marked rapid chromatographic immunoassay intended for the qualitative *in vitro* detection of both IgM and IgG antibodies to SARS-CoV-2 spike or nucleocapsid proteins in human serum, plasma or whole blood, for professional use in the laboratory and at point-of-care (26). A study utilizing real-world clinical samples in unvaccinated individuals demonstrated that the SARS-CoV-2 Rapid Antibody Test had comparable performance with the Elecsys Anti-SARS-CoV-2 assay that targets the nucleocapsid protein (27).

The objective of this study was to establish the performance of the SARS-CoV-2 Rapid Antibody Test and the Elecsys Anti-SARS-CoV-2 S assay in serum samples obtained from individuals who had received a COVID-19 vaccination.

## Materials and Methods

### Study design

The study was a retrospective performance study conducted at Roche Diagnostics GmbH (Mannheim, Germany). All samples were analyzed with the SARS-CoV-2 Rapid Antibody Test and compared with the Elecsys Anti-SARS-CoV-2 S assay. Tests were conducted as per manufacturer’s instructions (23).

### Samples

To assess antibodies following vaccination, a post-vaccination panel was prepared using 74 serum samples obtained from Boca Biolistics (Florida, USA), collected between February and April 2021, and stored at -20°C or colder (all available samples at the time of request). Serum samples were from individuals from the USA sampled prior to vaccination, after the first dose, or at least 14 days after the second dose of Moderna mRNA-1273 or Pfizer-BioNTech BNT162b2 (Supplemental Table 1).

Samples were separated into aliquots of 50 μL for the SARS-CoV-2 Rapid Antibody Test measurement and 200 μL for the Elecsys Anti-SARS-CoV-2 S reference measurement on the cobas^®^ 6000 and stored at -80°C until use.

An in-house panel of 15 negative serum samples, previously determined negative by Elecsys Anti-SARS-CoV-2 S assay, was utilized to assess the specificity of the SARS-CoV-2 Rapid Antibody Test. Samples were collected in June 2021 as part of an in-house blood donation service at Roche Diagnostics (Mannheim, Germany), anonymized, and stored at -80°C until use. No information about vaccination status was available. Evaluators were blinded as to the origin of the samples. Cross-reactivity to other pathogens was not analyzed in this study as this has been evaluated elsewhere (26, 27).

### SARS-CoV-2 Rapid Antibody Test

The SARS-CoV-2 Rapid Antibody Test detects the presence of SARS-CoV-2 IgM and IgG antibodies, and the intensity of the test line is dependent upon the level of SARS-CoV-2 antibodies present in the sample (26). In unvaccinated individuals, the reported sensitivity is 92.59% (7–14 days post-symptom onset [POS]) and 99.03% (>14 days POS), with specificity of 98.65% (26).

Each sample was tested with two different lots of the SARS-CoV-2 Rapid Antibody Test. The readout was visually assessed at 10 minutes by two different independent evaluators; these evaluators were not the operators who had performed the test. In accordance with manufacturer’s instructions, the sample was considered positive if the control line and at least one colored band/test line at G or M became apparent (qualitative determination). As an exploratory analysis, results were further defined by the intensity of the band to provide semi-quantitative data (Supplemental Table 2), with the signal intensity being rated according to a color scale. Data are presented separated by lot and by evaluator. In the event of an invalid result, the test would be repeated.

### Elecsys Anti-SARS-CoV-2 S assay

The Elecsys Anti-SARS-CoV-2 S assay results are reported automatically as the analyte concentration of each sample in U/mL, with <0.80 U/mL interpreted as negative and ≥0.80 U/mL interpreted as positive for anti-SARS-CoV-2 S antibodies (23). The reported sensitivity and specificity of this test 14 days post-PCR is 97.92% and 99.95%, respectively (24). The quantification range is between 0.4 and 250 U/mL and the reported limit of detection is 0.35 U/mL (23). If the sample concentration is >250 U/mL, the sample can be diluted up to 1:100; in these cases, the final result is determined as >25,000 U/mL (23).

In the event of an invalid result, the test would be repeated. All samples were measured in duplicate, and the result calculated as the arithmetic mean of both replicates. Assessors were blinded to the SARS-CoV-2 Rapid Antibody Test results and blinded as to the origin of the samples.

### Statistical analysis

The agreement rates comparing the SARS-CoV-2 Rapid Antibody Test with Elecsys Anti-SARS-CoV-2 S assay per vaccine type were calculated per lot. Two-sided 95% Clopper–Pearson confidence intervals (CI) for the agreement rates are given. Correlation between the semi-quantitative SARS-CoV-2 Rapid Antibody Test data and quantitative Elecsys Anti-SARS-CoV-2 S assay data was assessed using the Kendall’s rank correlation ⍰. Accuracy of the SARS-CoV-2 Rapid Antibody Test result — how often do the qualitative results agree with one another — was assessed by lot and evaluator. Statistical analyses were conducted using R (version 3.6.3) and JMP (version 15).

## Results

Overall, the study cohort consisted of 56 predominantly white/Caucasian individuals (median age 70 years; 55.36% female) with a variety of pre-existing medical conditions (Table 1). No invalid results were generated using either test.

**Table 1:**
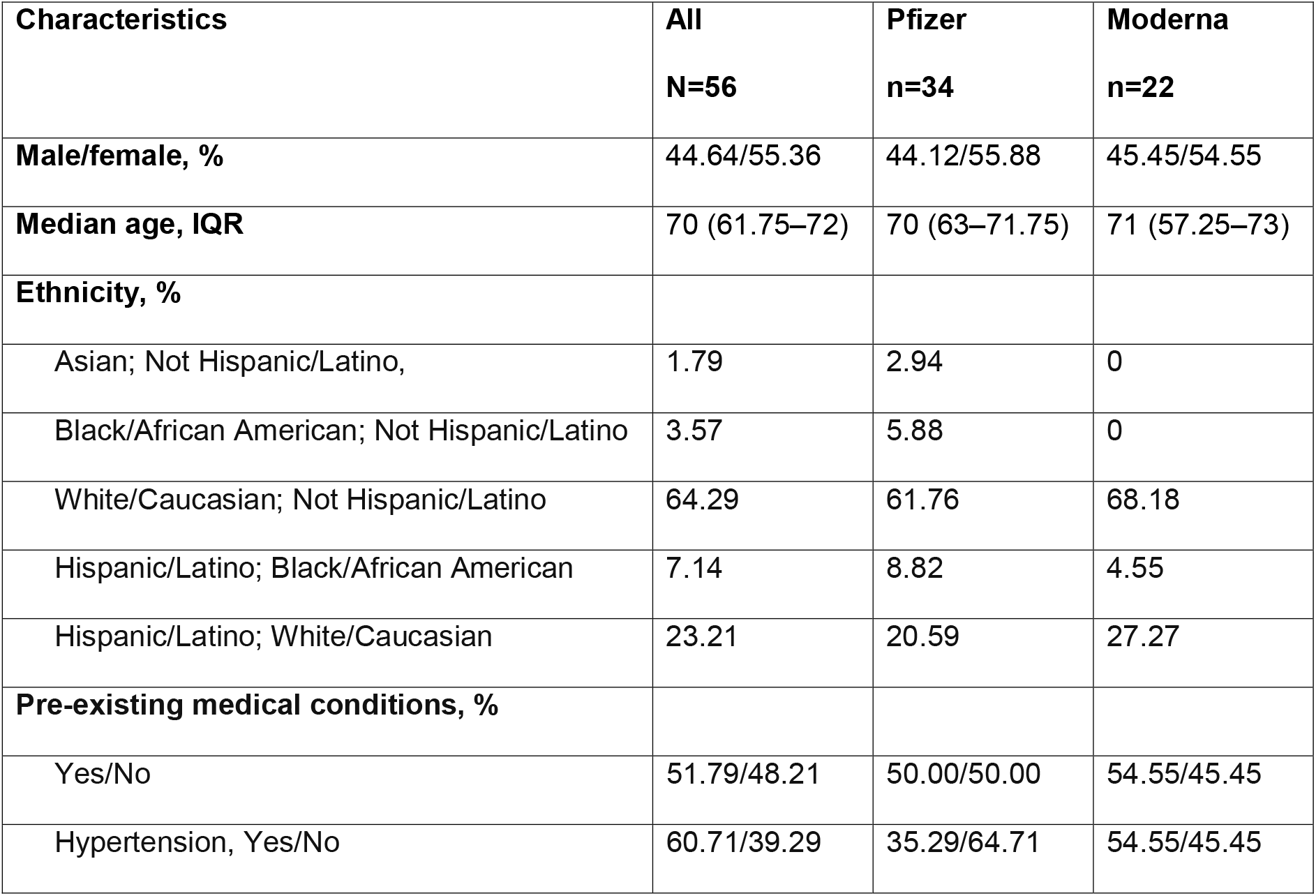
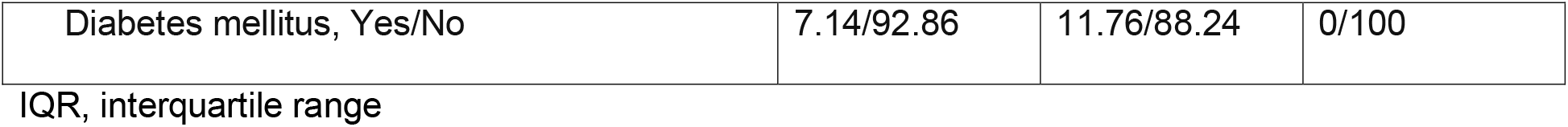
Demographics of the donors from the Boca Biolistics panel

| **Characteristics** | **All**<br>**N=56** | **Pfizer**<br>**n=34** | **Moderna**<br>**n=22** |
| --- | --- | --- | --- |
| **Male/female, %** | 44.64/55.36 | 44.12/55.88 | 45.45/54.55 |
| **Median age, IQR** | 70 (61.75–72) | 70 (63–71.75) | 71 (57.25–73) |
| **Ethnicity, %** |  |  |  |
| Asian; Not Hispanic/Latino, | 1.79 | 2.94 | 0 |
| Black/African American; Not Hispanic/Latino | 3.57 | 5.88 | 0 |
| White/Caucasian; Not Hispanic/Latino | 64.29 | 61.76 | 68.18 |
| Hispanic/Latino; Black/African American | 7.14 | 8.82 | 4.55 |
| Hispanic/Latino; White/Caucasian | 23.21 | 20.59 | 27.27 |
| **Pre-existing medical conditions, %** |  |  |  |
| Yes/No | 51.79/48.21 | 50.00/50.00 | 54.55/45.45 |
| Hypertension, Yes/No | 60.71/39.29 | 35.29/64.71 | 54.55/45.45 |
| Diabetes mellitus, Yes/No | 7.14/92.86 | 11.76/88.24 | 0/100 |

IQR, interquartile range

### Analyses of the SARS-CoV-2 Rapid Antibody Test (qualitative) with the Elecsys Anti-SARS-CoV-2 S assay (quantitative and qualitative)

Eight samples were obtained from patients who were sampled prior to vaccination. Whilst only two patients were reported to have been previously diagnosed with COVID-19, only one patient was IgG and IgM negative (by both the SARS-CoV-2 Rapid Antibody Test and Elecsys Anti-SARS-CoV-2 S assay). For the seven positive samples, Elecsys Anti-SARS-CoV-2 S total antibody titers ranged from 254.0 U/mL to >25,000 U/mL and all were identified as positive by the SARS-CoV-2 Rapid Antibody Test.

The post-vaccination antibody analysis included 25 samples from individuals vaccinated with mRNA-1273 and 42 vaccinated with BNT162b2. The dosing interval between first and second vaccine doses for BNT162b2 was a median of 21 days (minimum 19 days, maximum 23 days) and a median of 28 days (minimum 21 days, maximum 32 days) for mRNA-1273. There was good agreement between the SARS-CoV-2 Rapid Antibody Test (IgG) and Elecsys Anti-SARS-CoV-2 S qualitative measurements at the different assessment points for both the Moderna and Pfizer-BioNTech vaccines ranging from 97.06% (95% CI 84.67, 99.93) to 100% (95% CI 89.72, 100) (Table 2). There was one false-negative sample with evaluator 2 (after second dose of BNT162b2, lot 1 and lot 2), however this sample was classified as positive by evaluator 1.

**Table 2:** Agreement between SARS-CoV-2 Rapid Antibody Test (IgG and IgM) and Elecsys Anti-SARS-CoV-2 S assay (reference test) qualitative measurements as measured by lot and evaluator using the Boca Biolistics donor panel, and after one dose or at least 14 days after two doses of Moderna mRNA-1273 or Pfizer-BioNTech BNT162b2

|  |  | mRNA-1273 |  |  |  |  |  |  |  | BNT162b2 |  |  |  |  |  |  |  |
| --- | --- | --- | --- | --- | --- | --- | --- | --- | --- | --- | --- | --- | --- | --- | --- | --- | --- |
|  |  | 1 dose |  |  |  | 2 doses |  |  |  | 1 dose |  |  |  | 2 doses |  |  |  |
|  |  | Evaluator 1 |  | Evaluator 2 |  | Evaluator 1 |  | Evaluator 2 |  | Evaluator 1 |  | Evaluator 2 |  | Evaluator 1 |  | Evaluator 2 |  |
|  |  | Lot 1 | Lot 2 | Lot 1 | Lot 2 | Lot 1 | Lot 2 | Lot 1 | Lot 2 | Lot 1 | Lot 2 | Lot 1 | Lot 2 | Lot 1 | Lot 2 | Lot 1 | Lot 2 |
| **Ig G** | **N** | 5 | 5 | 5 | 5 | 19 | 19 | 19 | 19 | 8 | 8 | 8 | 8 | 34 | 34 | 34 | 34 |
|  | **N +** | 5 | 5 | 5 | 5 | 19 | 19 | 19 | 19 | 7 | 7 | 7 | 7 | 34 | 34 | 34 | 34 |
|  | **N -** | - | - | - | - | - | - | - | - | 1* | 1* | 1* | 1* | - | - | - | - |
|  | **TP** | 5 | 5 | 5 | 5 | 19 | 19 | 19 | 19 | 7 | 7 | 7 | 7 | 34 | 34 | 33 | 33 |
|  | **FN** | 0 | 0 | 0 | 0 | 0 | 0 | 0 | 0 | 0 | 0 | 0 | 0 | 0 | 0 | 1 | 1 |
|  | **PPA, %** | 100 | 100 | 100 | 100 | 100 | 100 | 100 | 100 | 100 | 100 | 100 | 100 | 100 | 100 | 97.0<br>6 | 97.0<br>6 |
|  | **lower** | 47.82 | 47.82 | 47.82 | 47.82 | 82.3<br>5 | 82.3<br>5 | 82.3<br>5 | 82.3<br>5 | 59.0<br>4 | 59.0<br>4 | 59.0<br>4 | 59.0<br>4 | 89.7<br>2 | 89.7<br>2 | 84.6<br>7 | 84.6<br>7 |
|  | **CI** |  |  |  |  |  |  |  |  |  |  |  |  |  |  |  |  |
|  | **upper CI** | 100 | 100 | 100 | 100 | 100 | 100 | 100 | 100 | 100 | 100 | 100 | 100 | 100 | 100 | 99.93 | 99.93 |
| **Ig M** | **N** | 5 | 5 | 5 | 5 | 19 | 19 | 19 | 19 | 8 | 8 | 8 | 8 | 34 | 34 | 34 | 34 |
|  | **N +** | 5 | 5 | 5 | 5 | 19 | 19 | 19 | 19 | 7 | 7 | 7 | 7 | 34 | 34 | 34 | 34 |
|  | **N -** | - | - | - | - | - | - | - | - | 1* | 1* | 1* | 1* | - | - | - | - |
|  | **TP** | 1 | 1 | 4 | 3 | 4 | 4 | 6 | 4 | 2 | 2 | 3 | 2 | 6 | 6 | 5 | 5 |
|  | **FN** | 4 | 4 | 1 | 2 | 15 | 15 | 13 | 15 | 5 | 5 | 4 | 5 | 28 | 28 | 29 | 29 |
|  | **PPA, %** | 20.00 | 20.00 | 80.00 | 60.00 | 21.05 | 21.05 | 31.58 | 21.05 | 28.57 | 28.57 | 42.86 | 28.57 | 17.65 | 17.65 | 14.71 | 14.71 |
|  | **lower CI** | 0.51 | 0.51 | 28.36 | 14.66 | 6.05 | 6.05 | 12.58 | 6.05 | 3.67 | 3.67 | 9.9 | 3.67 | 6.76 | 6.76 | 4.95 | 4.95 |
|  | **upper CI** | 71.64 | 71.64 | 99.49 | 94.73 | 45.57 | 45.57 | 56.55 | 45.57 | 70.96 | 70.96 | 81.59 | 70.96 | 34.53 | 34.53 | 31.06 | 31.06 |

\*1 sample was detected as antibody-negative by the Elecsys Anti-SARS-CoV-2 S and the SARS-CoV-2 Rapid Antibody Test.
+, positive by the reference test (Elecsys Anti-SARS-CoV-2 S assay); -, negative by the reference test (Elecsys Anti-SARS-CoV-2
S assay); CI, 95% confidence intervals; TP, true-positive; FN, false-negative; PPA, positive percent agreement

There was poor agreement between the SARS-CoV-2 Rapid Antibody Test (IgM) and Elecsys Anti-SARS-CoV-2 S assay qualitative measurements at the different assessment points for both BNT162b2 and mRNA-1273 ranging from 14.71% (95% CI 4.95, 31.06) to 80.00% (95% CI 28.36, 99.49) with many false-negatives (Table 2).

Using the in-house negative panel, there were no false-positive results for either lot or evaluator (**Error! Reference source not found**.).

### Analyses of the SARS-CoV-2 Rapid Antibody Test (semi-quantitative) with the Elecsys Anti-SARS-CoV-2 S assay (quantitative)

There was a medium-to-strong correlation between the semi-quantitative SARS-CoV-2 Rapid Antibody Test (IgG) and quantitative Elecsys Anti-SARS-CoV-2 S assay in samples taken after both doses of the vaccines (Figure 1), with higher intensity bands being associated with higher total anti-S antibody titer (with Kendall’s ⍰ up to 0.599 for IgG mRNA-1273 [p=0.0019] and 0.6833 [p<0.0001] for IgG BNT162b2) (**Error! Reference source not found**.). There was no correlation between the quantitative Elecsys Anti-SARS-CoV-2 S assay result and the intensity of the SARS-CoV-2 Rapid Antibody Test (IgM) (**Error! Reference source not found**.).

**Figure 1:**
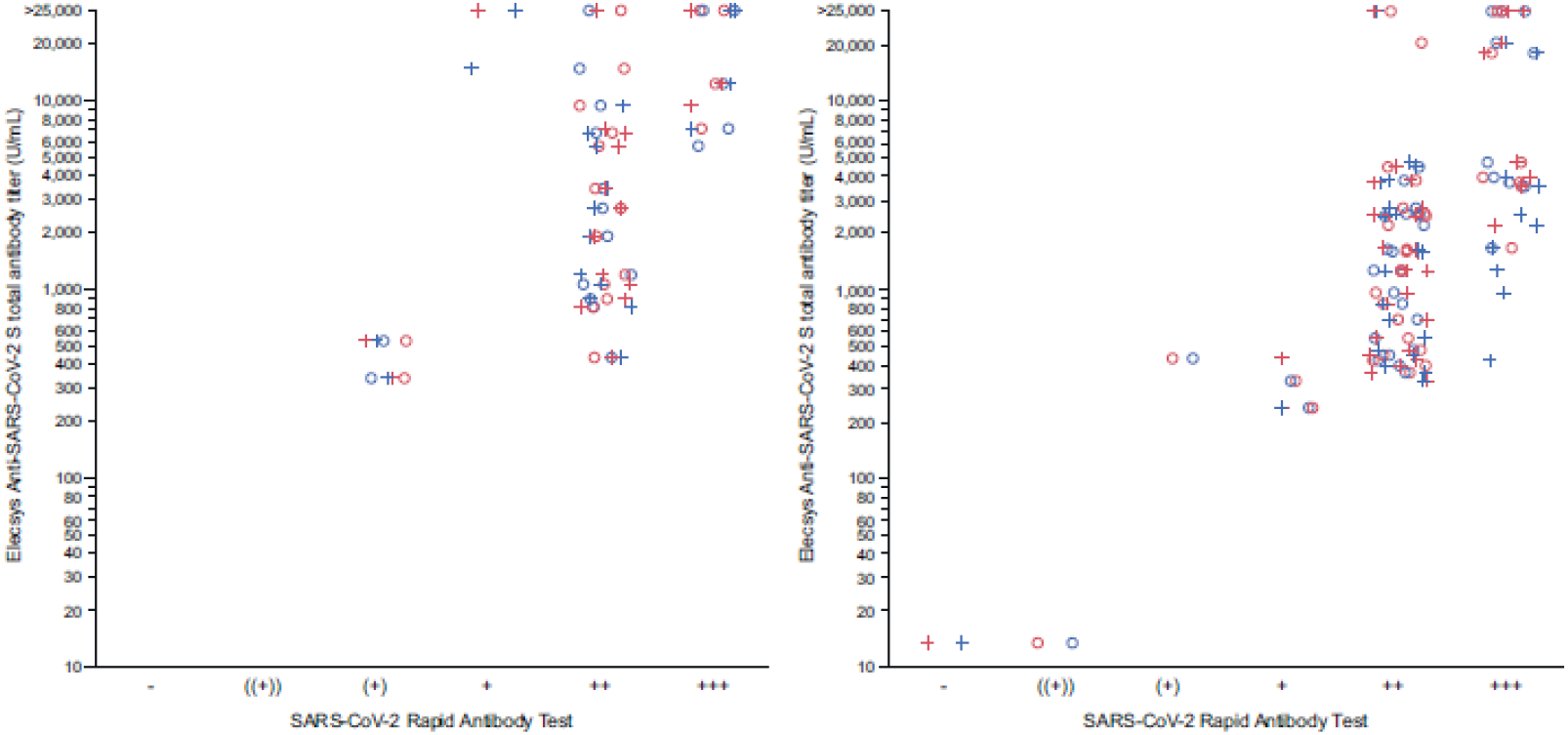
SARS-CoV-2 Rapid Antibody Test semi-quantitative IgG vs Elecsys Anti-SARS-CoV-2 S assay total antibody titer following vaccination with Moderna mRNA-1273 (left panel) or Pfizer-BioNTech BNT162b2 (right panel) at least 14 days following second dose Results from the SARS-CoV-2 Rapid Antibody Test were classified as negative or one of the five levels of increasing positivity based upon line intensity (((+)), (+), +, ++ or +++). Maximum value attainable by the Elecsys Anti-SARS-CoV-2 assay was 25,000 U/mL. Higher values are indicated as >25,000 U/mL. Blue = Lot 1; red = Lot 2; ○ = Evaluator 1; + = Evaluator 2.

Agreement between the semi-quantitative SARS-CoV-2 Rapid Antibody Test (IgM) and quantitative Elecsys Anti-SARS-CoV-2 S assay was poor, with many samples designated as negative on the SARS-CoV-2 Rapid Antibody Test (IgM) even in the presence of high anti-S antibody titers (Results from the SARS-CoV-2 Rapid Antibody Test were classified as negative or one of the five levels of increasing positivity based upon line intensity (((+)), (+), +, ++ or +++). Maximum value attainable by the Elecsys Anti-SARS-CoV-2 assay was 25,000 U/mL. Higher values are indicated as >25,000 U/mL. Blue = Lot 1; red = Lot 2; ○ = Evaluator 1; + = Evaluator 2.

Figure 2).

**Figure 2:**
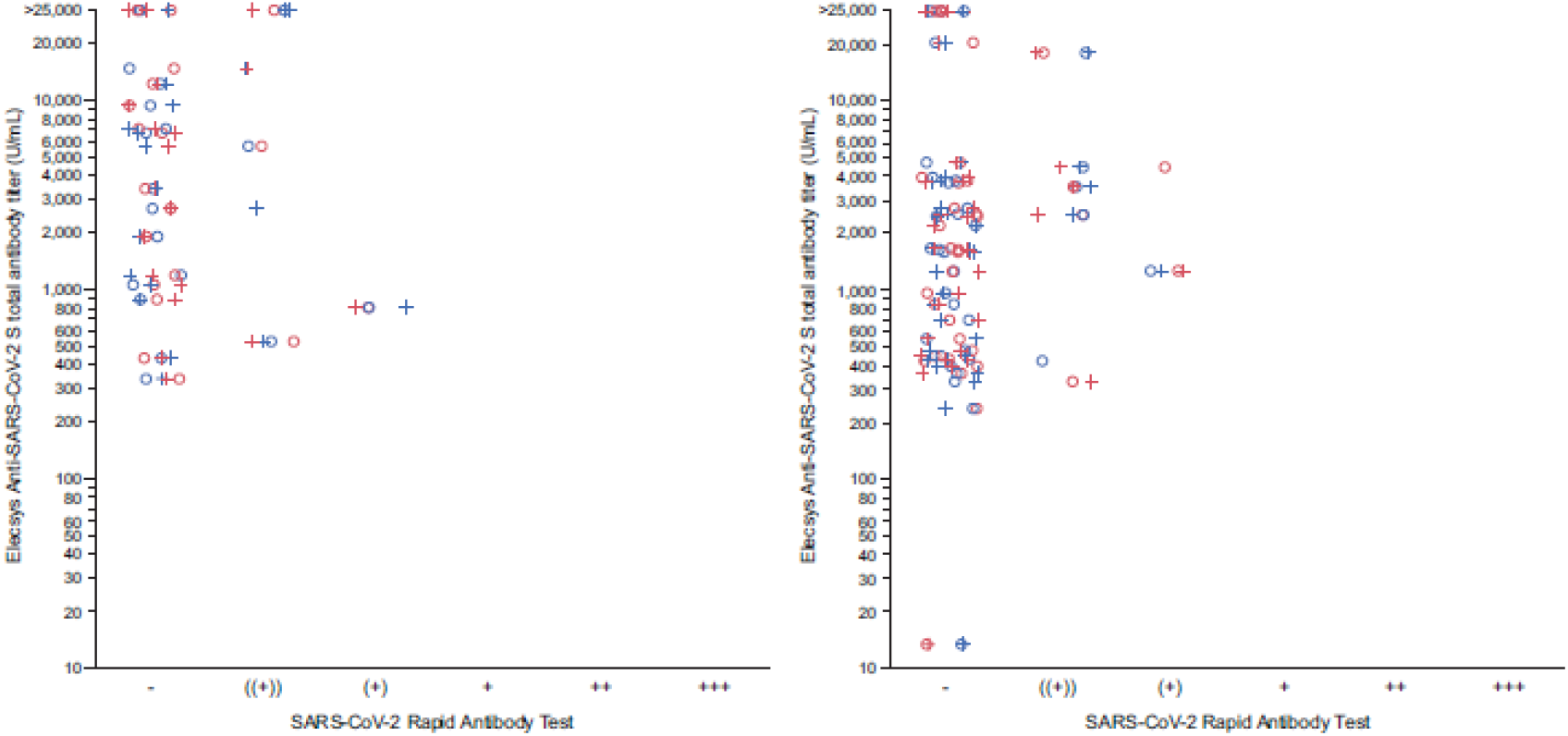
SARS-CoV-2 Rapid Antibody Test semi-quantitative IgM vs Elecsys Anti-SARS-CoV-2 S assay total antibody titer following vaccination with Moderna mRNA-1273 (left panel) or Pfizer-BioNTech BNT162b2 (right panel) at least 14 days following second dose Results from the SARS-CoV-2 Rapid Antibody Test were classified as negative or one of the five levels of increasing positivity based upon line intensity (((+)), (+), +, ++ or +++). Maximum value attainable by the Elecsys Anti-SARS-CoV-2 assay was 25,000 U/mL. Higher values are indicated as >25,000 U/mL. Blue = Lot 1; red = Lot 2; ○ = Evaluator 1; + = Evaluator 2.

### Accuracy of the SARS-CoV-2 Rapid Antibody Test

Comparison of the SARS-CoV-2 Rapid Antibody Test (IgG) results by lot or evaluator revealed that these factors did not influence the results, with a minimum accuracy estimate of 97.06% (**Error! Reference source not found**.).

SARS-CoV-2 Rapid Antibody Test (IgM) positivity appeared to be influenced by evaluator, with a minimum accuracy estimate of 40% and considerable variability in accuracy estimates across the different time points and vaccines (**Error! Reference source not found**.).

### Evaluation of Elecsys Anti-SARS-CoV-2 S assay titer post-vaccination

Antibody titers by days following final vaccination are shown in Figure 3. Following complete vaccination with BNT162b2 or mRNA-1273 and up to approximately 20 days, there was significant variability with both high (maximum >25,000 U/mL) and low (minimum 238.2 U/mL for BNT162b2 and 340.2 U/mL for mRNA-1273) anti-S titers. After 30 days, anti-S titers for both vaccines commonly declined to below 5,000 U/mL (overall minimum 13.53 U/mL 25 days after vaccination with BNT162b2).

**Figure 3:**
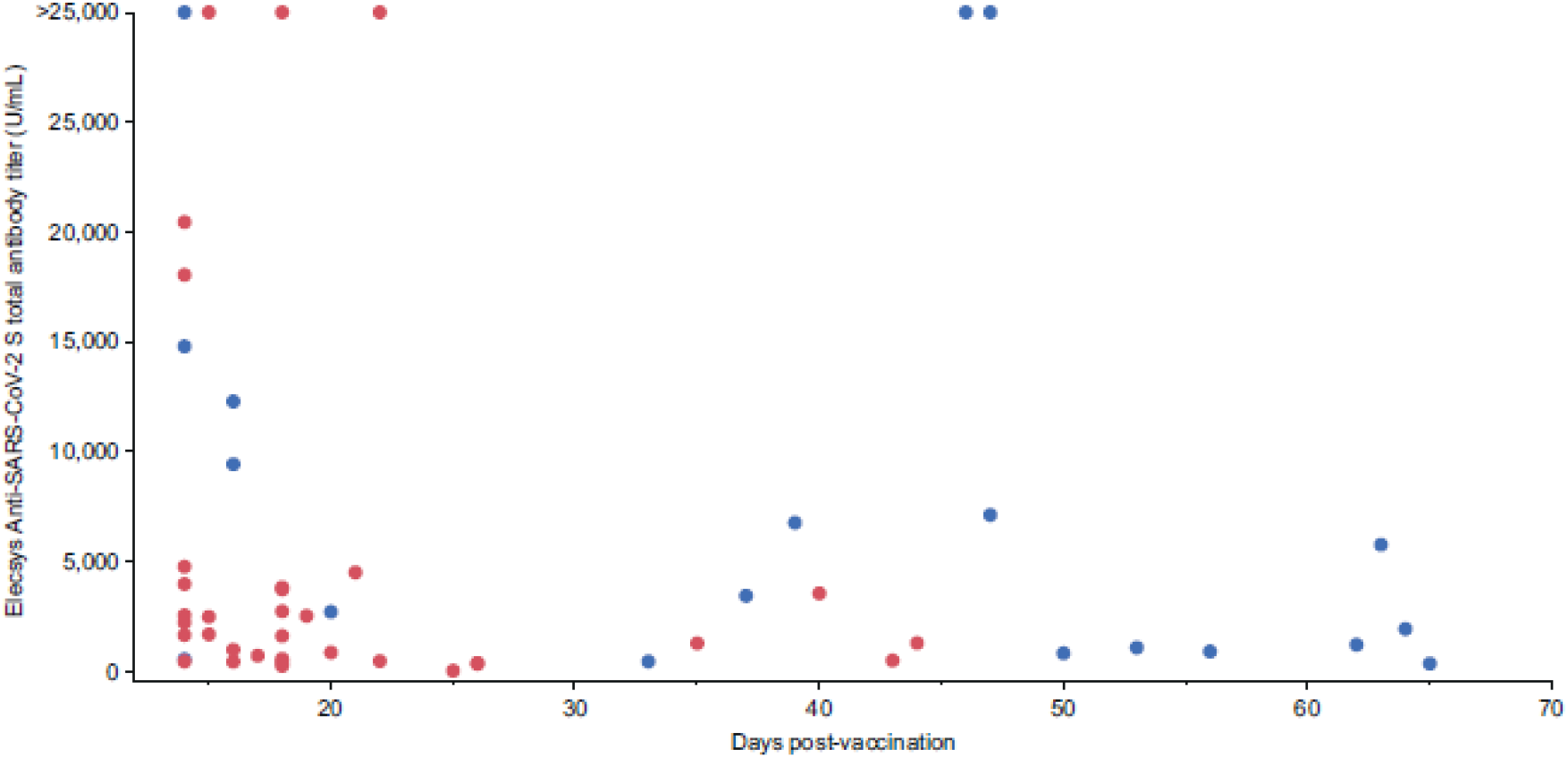
Elecsys Anti-SARS-CoV-2 S assay total antibody titer over time following vaccination with Moderna mRNA-1273 or Pfizer-BioNTech BNT162b2 (second dose) Maximum value attainable by the Elecsys Anti-SARS-CoV-2 assay was 25,000 U/mL. Higher values are indicated as >25,000 U/mL. Blue = mRNA-1273; red = BNT162b2

The median time between final vaccination and final measurement for those vaccinated with BNT162b2 was 18 days (minimum 14 days, maximum 44 days) and 46 days (minimum 14 days, maximum 65 days) for mRNA-1273. Anti-S antibody titers were numerically higher in participants vaccinated mRNA-1273 (median 2,307 U/mL; interquartile range [IQR] 872–6,845) than in those who received BNT162b2 (median 1,601 U/mL; IQR 467–3,131).

### Longitudinal analysis

A small number of individuals had samples available from all three time points (BNT162b2 cohort), allowing longitudinal evaluation of anti-S antibody titer levels; all except one showed high levels of anti-S antibody titers at the first time point (prior to vaccination). There was considerable variance in the effect of vaccination upon anti-S antibody titers, however the greatest change was for the individual who was infection-naïve (<0.4 U/mL to 1,645 U/mL following the second dose). For five out of eight individuals, including those with previously diagnosed COVID-19, anti-S antibody titers decreased over time despite vaccination with BNT162b2 (**Error! Reference source not found**.).

## Discussion

Serologic assays are an essential tool for quality control of vaccines and to determine the response to vaccination. Although a correlate of immunity has not yet been established for COVID-19 vaccines, antibody titers after natural infection and vaccination have been associated with protection from symptomatic SARS-CoV-2 infection (28); and rapid point-of-care assays can potentially be of use in this setting with particular advantages over centralized testing, such as speed and ease of use.

We evaluated the use of the SARS-CoV-2 Rapid Antibody Test in vaccinated individuals in comparison with the Elecsys Anti-SARS-CoV-2 S assay.

Our results indicate that the Elecsys Anti-SARS-CoV-2 S assay detects anti-S antibodies after the first dose of both BNT162b2 and mRNA-1273, in agreement with previous studies of the adaptive humoral response to vaccination in various populations (21, 22, 29–33).

Whilst the performance of rapid antibody tests has been reported in unvaccinated individuals (34–36), no studies have yet reported upon their use in vaccinated individuals. The SARS-CoV-2 Rapid Antibody Test (IgG) compared favorably to the Elecsys Anti-SARS-CoV-2 S assay with agreement rates above 97.06%, after one dose and two doses, similar to what has been observed in unvaccinated individuals (27). Assessment of the influence of lot or evaluator revealed that agreement of the SARS-CoV-2 Rapid Antibody Test (IgG) with the Elecsys Anti-SARS-CoV-2 S assay was not highly influenced by either factor, suggesting that the SARS-CoV-2 Rapid Antibody Test (IgG) is robust under normal usage conditions.

There was low agreement of the SARS-CoV-2 Rapid Antibody Test (IgM) with the Elecsys Anti-SARS-CoV-2 S assay because, in general, there is a lower sensitivity for IgM compared with IgG for point-of-care lateral flow assays, as exemplified in a recent analysis of healthcare workers vaccinated with the first dose of the Pfizer vaccine and sampled 21–24 days thereafter (37). In addition, the Elecsys Anti-SARS-CoV-2 S assay preferentially detects high-affinity IgG antibodies that compete with IgM antibodies (23), contributing to the lower agreement rates with IgM results. In agreement with previous research on the use of lateral flow assays in unvaccinated individuals (38, 39), we also report lower evaluator concordance when reading IgM compared with IgG bands. Jones and colleagues found that IgM bands are often weak positives (39), consistent with our data in vaccinated individuals, potentially resulting in an increased likelihood of a miscall and contributing to the low reader agreement.

The vaccines were administered in accordance with the relevant primary dosing schedules, with approximately 3 weeks for BNT162b2 and 1 month for mRNA-1273 between doses (17, 18). Whilst the data reported herein suggest some variation in the level of antibody titers afforded by the vaccines, two doses of either mRNA-1273 or BNT162b2 induced high levels of anti-S antibodies. Anti-S total antibody titers were higher in participants vaccinated with mRNA-1273 than BNT162b2, in accordance with previous publications (40). Whilst the clinical significance of quantitative differences in antibody titers following COVIDl7l19 vaccination have not yet been established, understanding the differences in antibody titer can support individual choices and assist with policy decisions.

The longitudinal analysis revealed that five individuals in the BNT162b2 cohort appeared to have previously undiagnosed SARS-CoV-2 infection prior to vaccination, and the effect of vaccination upon anti-S antibody titers in this group at the individual level was highly variable (relative decrease or increase after the first dose). Overall, after primary vaccination with two doses, all except one of those with previous natural infection had higher anti-S antibody titers than the infection-naïve participant, in agreement with previous reports which support higher titers following vaccination in previously infected individuals (40–42).

The findings of this study are subject to several limitations. The performance of the SARS-CoV-2 Rapid Antibody Test in vaccinated individuals was based on the evaluation of a relatively small panel of samples and utilized only two vaccines. However, the cohort tested was generally representative of an aged US population, with hypertension and diabetes mellitus in line with age-related expectations of prevalence (43, 44). Whilst the median anti-S antibody titers induced by vaccination differed between those vaccinated with BNT162b2 or mRNA-1273, the majority of the post-vaccination BNT162b2 samples were collected earlier than samples collected for mRNA-1273, there were more data points for BNT162b2, and the data for mRNA-1273 samples spanned a longer time frame (last sample was post 60 days for mRNA-1273 compared with <45 days for BNT162b2). Furthermore, most of the samples used in the longitudinal analysis were from positive pre-vaccinated individuals, which limits our findings.

The clinical relevance of semil7lquantitative results is currently unknown and, at present, cannot be interpreted as an indication of varying levels of immunity. In addition, semi-quantitative evaluation of the intensity of the bands is determined by the reader and therefore subjective, even with the use of a color scale guide and training. The performance of the SARS-CoV-2 Rapid Antibody Test seen herein can also not be extrapolated to immunocompromised individuals whose immune response post vaccination may be more variable (45). However, semi-quantitative SARS-CoV-2 Rapid Antibody Test (IgG) and quantitative Elecsys Anti-SARS-CoV-2 S assay results correlated well, suggesting that SARS-CoV-2 Rapid Antibody Test (IgG) is helpful in understanding the immune response post vaccination, particularly in the absence of centralized and fully automated testing systems. The current data support the use of the SARS-CoV-2 Rapid Antibody Test (IgG) in the vaccinated population.

## Data Availability

Data available upon request. For more information on the study and data sharing, qualified researchers may contact the corresponding author, Eva Urlaub.

## Ethical approval and consent

Anonymized samples were provided by the commercial supplier Boca Biolistics and collected under their ethical approval system. Ethical approval for the in-house panel was provided by Ethik-Kommission II der Universität Heidelberg, Medizinische Fakultät, Mannheim, Germany. Samples were anonymized, and collected and tested in accordance with applicable regulations, including relevant European Union directives and regulations, and the principles of the Declaration of Helsinki.

## Acknowledgements

Medical writing support was provided by Corrinne Segal, Elements Communications Ltd, Westerham, UK. The authors would like to thank Rabea Held, Regina Draude, Maureen Adamietz, Shervan Horo, and Jlenia Marconi of Roche Diagnostics for their contribution to the study. COBAS, COBAS E, and ELECSYS are trademarks of Roche. All other product names and trademarks are the property of their respective owners’. The specific application (use in vaccinated individuals) is currently outside of the intended use of the SARS-CoV-2 Rapid Antibody Test. The Elecsys Anti-SARS-CoV-2 S assay is approved under an Emergency Use Authorization in the US.

