## Supplemental for "Evaluation of the Roche SARS-CoV-2 Rapid Antibody Test in samples from vaccinated individuals"

Supplemental Table 1: Boca Biolistics sample panel

| | Sampled prior to vaccination | Sampled after first dose | Sampled ≥ 14 days after second dose | Sub-total |
|---|---|---|---|---|
| **Moderna** | 8* | 5† | 19 | 24 |
| **Pfizer-BioNTech** | | 8 | 34 | 42 |
| **Sub-total** | | 13 | 53 | 66 |
| **Total samples** | 74 | | | |

*2 samples came from donors who were previously diagnosed with COVID-19 (known infection, unknown whether diagnosis was PCR-confirmed). Diagnosis was 3 months prior to the date of the first draw for one individual and 6 months for the second individual

†Duplicate samples from the same donor, taken at the same visit, were available – only the first sample was used in analyses

Supplemental Table 2: Interpretation of SARS-CoV-2 Rapid Antibody Test for semi-quantitative analysis

| Result | Symbol* | Description |
|---|---|---|
| **IgM positive** | ((+)), (+), +, ++, +++ | • Control line and IgM signal line both visible<br>• Faint signal lines are rated as positive<br>• Signal intensity will be assessed using the color scale |
| **IgG positive** | ((+)), (+), +, ++, +++ | • Control line and IgG signal line both visible<br>• Faint signal lines are rated as positive<br>• Signal intensity will be assessed using the color scale |
| **Negative** | - | • Control line visible<br>• No signal line |
| **Invalid** | / | • No control line<br>• Unusual background of the test strip<br>• Further reasons |

*Daylight lamps were used to assist the visual interpretation of the bands.

Results from the SARS-CoV-2 Rapid Antibody Test were classified as invalid, negative or one of the five levels of increasing positivity based upon line intensity (((+)), (+), +, ++ or +++)

Supplemental Table 3: Agreement between SARS-CoV-2 Rapid Antibody Test (IgG and IgM) and Elecsys Anti-SARS-CoV-2 S assay (reference test) qualitative measurements, as measured by lot and evaluator for in-house negative panel

| | | **Evaluator 1** | | **Evaluator 2** | |
|---|---|---|---|---|---|
| | | **Lot 1** | **Lot 2** | **Lot 1** | **Lot 2** |
| **IgG** | **N** | 15 | 15 | 15 | 15 |
| | **N -** | 15 | 15 | 15 | 15 |
| | **FP** | 0 | 0 | 0 | 0 |
| | **TN** | 15 | 15 | 15 | 15 |
| | ***NPA, %*** | *100* | *100* | *100* | *100* |
| | ***lower CI*** | *78.2* | *78.2* | *78.2* | *78.2* |
| | ***upper CI*** | *100* | *100* | *100* | *100* |
| **IgM** | **N** | 15 | 15 | 15 | 15 |
| | **N -** | 15 | 15 | 15 | 15 |
| | **FP** | 0 | 0 | 0 | 0 |
| | **TN** | 15 | 15 | 15 | 15 |
| | ***NPA, %*** | *100* | *100* | *100* | *100* |
| | ***lower CI*** | *78.2* | *78.2* | *78.2* | *78.2* |
| | ***upper CI*** | *100* | *100* | *100* | *100* |

-, negative by the reference test (Elecsys Anti-SARS-CoV-2 S assay); CI, 95% confidence intervals; FP, false-positive; TN, true-negative; NPA, negative percent agreement

Supplemental Table 4. Kendall’s correlation between the SARS-CoV-2 Rapid Antibody Test semi-quantitative results and the Elecsys Anti-SARS-CoV-2 S total antibody titer for samples from individuals vaccinated with Moderna mRNA-1273 or Pfizer-BioNTech BNT162b2 (the analysis includes only those who have had two doses). Moderna: n=19 for each lot/evaluator. Pfizer: n=34 for each lot/evaluator. Significant p-values indicated in red.

| | | Kendall's τ | p-value |
|---|---|---|---|
| **mRNA-1273** | | | |
| **IgM** | | | |
| Elecsys_Quant | Rapid AB test, Lot 1 Evaluator 1 | -0.0875 | 0.6541 |
| Elecsys_Quant | Rapid AB test, Lot 1 Evaluator 2 | 0.1101 | 0.5722 |
| Elecsys_Quant | Rapid AB test, Lot 2 Evaluator 1 | -0.0875 | 0.6541 |
| Elecsys_Quant | Rapid AB test, Lot 2 Evaluator 2 | 0.0097 | 0.9603 |
| **IgG** | | | |
| Elecsys_Quant | Rapid AB test, Lot 1 Evaluator 1 | 0.5729 | 0.0029 |
| Elecsys_Quant | Rapid AB test, Lot 1 Evaluator 2 | 0.3712 | 0.0479 |
| Elecsys_Quant | Rapid AB test, Lot 2 Evaluator 1 | 0.599 | 0.0019 |
| Elecsys_Quant | Rapid AB test, Lot 2 Evaluator 2 | 0.3009 | 0.1103 |
| **BNT162b2** | | | |
| **IgM** | | | |
| Elecsys_Quant | Rapid AB test, Lot 1 Evaluator 1 | 0.1126 | 0.4304 |
| Elecsys_Quant | Rapid AB test, Lot 1 Evaluator 2 | 0.1977 | 0.1667 |
| Elecsys_Quant | Rapid AB test, Lot 2 Evaluator 1 | 0.1021 | 0.4718 |
| Elecsys_Quant | Rapid AB test, Lot 2 Evaluator 2 | 0.0933 | 0.5136 |

| IgG | | | |
|---|---|---|---|
| Elecsys_Quant | Rapid AB test, Lot 1 Evaluator 1 | 0.6833 | <0.0001 |
| Elecsys_Quant | Rapid AB test, Lot 1 Evaluator 2 | 0.4075 | 0.0035 |
| Elecsys_Quant | Rapid AB test, Lot 2 Evaluator 1 | 0.5881 | <0.0001 |
| Elecsys_Quant | Rapid AB test, Lot 2 Evaluator 2 | 0.5956 | <0.0001 |

Elecsys, Elecsys Anti-SARS-CoV-2 S assay; Quant, quantitative; Rapid AB test, SARS-CoV-2 Rapid Antibody Test

Supplemental Figure 1: Forest plot showing accuracy for SARS-CoV-2 Rapid Antibody Test (IgG), lot-to-lot and evaluator-to-evaluator, after vaccination with Moderna mRNA-1273 or Pfizer-BioNTech BNT162b2.

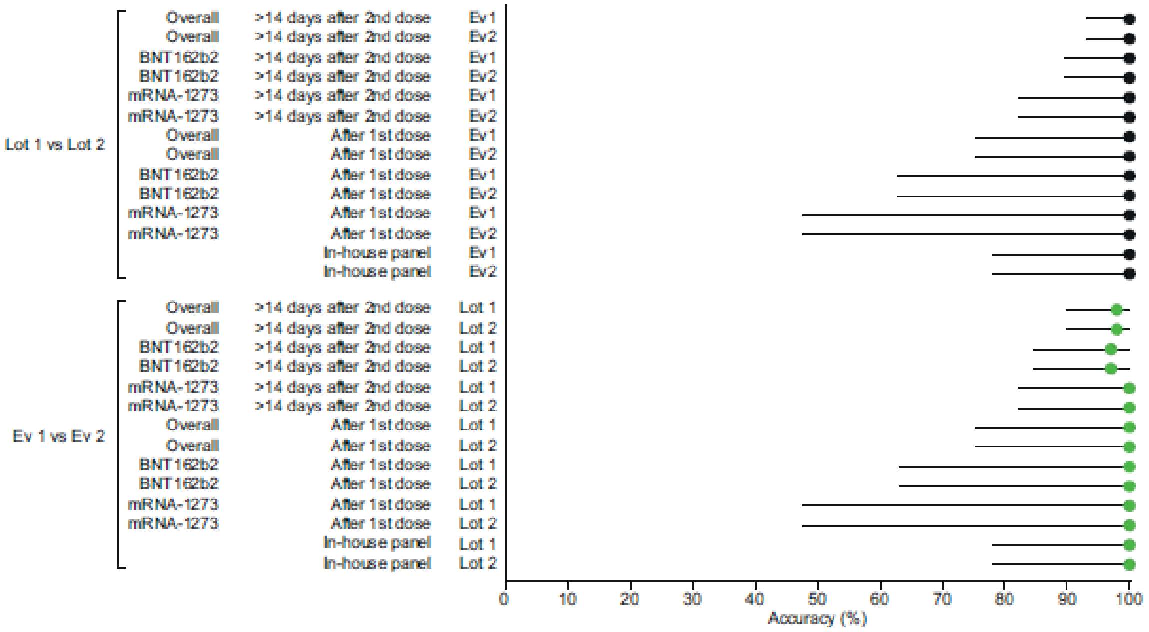


The horizontal line represents the 95% confidence intervals of the point estimate.

Overall represents the combined accuracy data for mRNA-1273 and BNT162b2

Ev, evaluator

Supplemental Figure 2: Forest plot showing accuracy estimates for SARS-CoV-2 Rapid Antibody Test (IgM), lot-to-lot and evaluator-to-evaluator, after vaccination with Moderna mRNA-1273 of Pfizer-BioNTech BNT162b2.

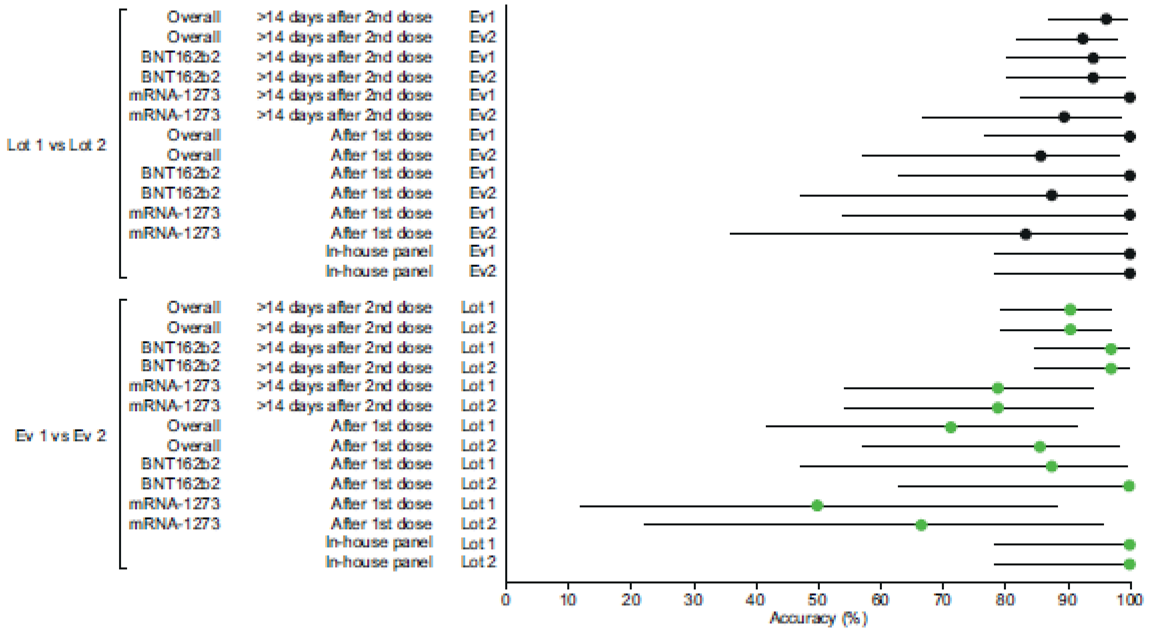


The horizontal line represents the 95% confidence intervals of the point estimate.
Overall represents the combined accuracy data for mRNA-1273 and BNT162b2

Ev, evaluator

Supplemental Figure 3: Longitudinal analysis of Elecsys Anti-SARS-CoV-2 S assay total antibody titers in individuals with serial samples available after vaccination with Pfizer-BioNTech BNT162b2

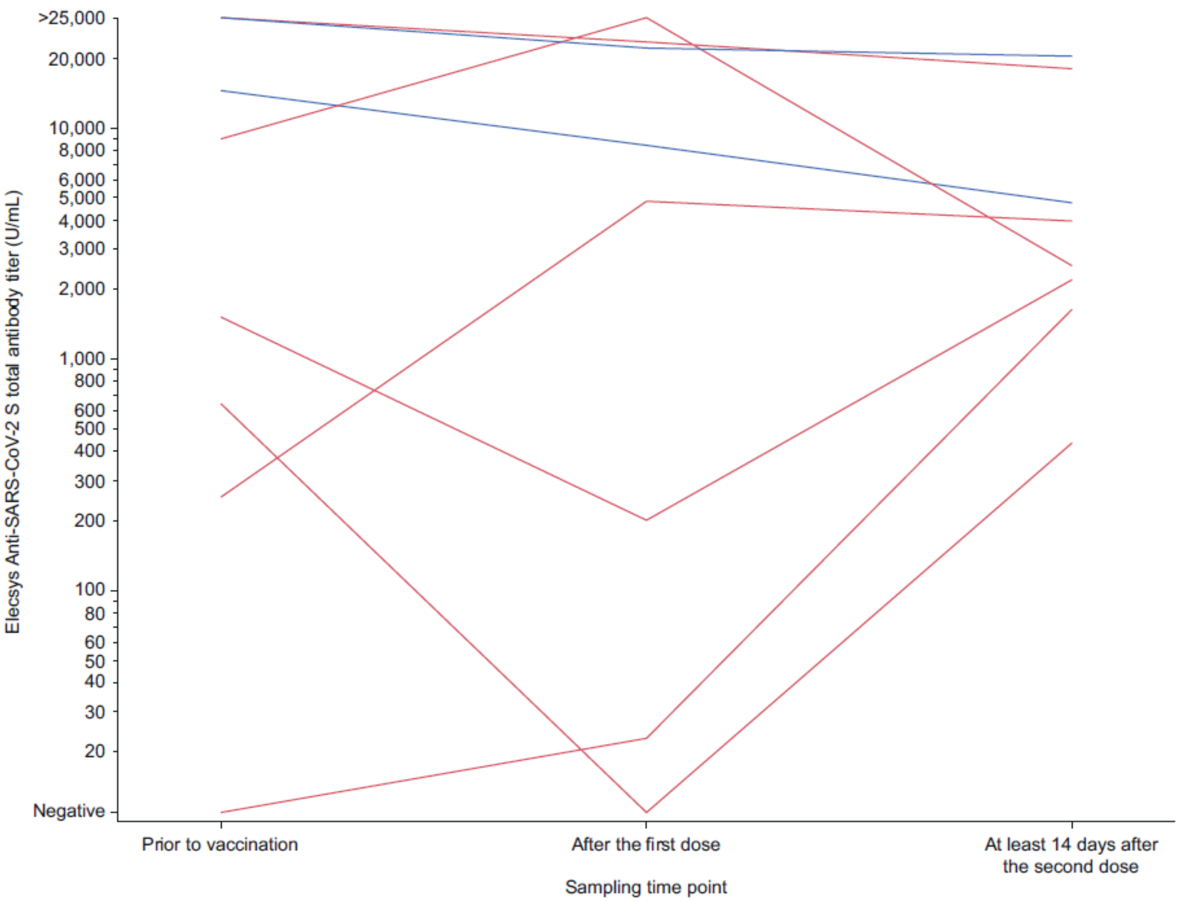


Blue color represents those individuals diagnosed with COVID-19 prior to vaccination

### STARD checklist

| Section & Topic | No | Item | Reported on page # |
|---|---|---|---|
| **TITLE OR ABSTRACT** | | | |
| | **1** | Identification as a study of diagnostic accuracy using at least one measure of accuracy (such as sensitivity, specificity, predictive values, or AUC) | 2 (agreement) |
| **ABSTRACT** | | | |
| | **2** | Structured summary of study design, methods, results, and conclusions (for specific guidance, see STARD for Abstracts) | 1/2 |
| **INTRODUCTION** | | | |
| | **3** | Scientific and clinical background, including the intended use and clinical role of the index test | 4/5 |
| | **4** | Study objectives and hypotheses | 5 |
| **METHODS** | | | |
| *Study design* | **5** | Whether data collection was planned before the index test and reference standard were performed (prospective study) or after (retrospective study) | 5 |
| *Participants* | **6** | Eligibility criteria | 6 |
| | **7** | On what basis potentially eligible participants were identified (such as symptoms, results from previous tests, inclusion in registry) | 6 |
| | **8** | Where and when potentially eligible participants were identified (setting, location and dates) | 6 |
| | **9** | Whether participants formed a consecutive, random or convenience series | NA |
| *Test methods* | **10a** | Index test, in sufficient detail to allow replication | 7 + Supplemental Table 2 |
| | **10b** | Reference standard, in sufficient detail to allow replication | 7 |
| | **11** | Rationale for choosing the reference standard (if alternatives exist) | NA |
| | **12a** | Definition of and rationale for test positivity cut-offs or result categories of the index test, distinguishing pre-specified from exploratory | 7 |
| | **12b** | Definition of and rationale for test positivity cut-offs or result categories of the reference standard, distinguishing pre-specified from exploratory | 7 |
| | **13a** | Whether clinical information and reference standard results were available to the performers/readers of the index test | 6/8 |
| | **13b** | Whether clinical information and index test results were available to the assessors of the reference standard | 6/8 |
| *Analysis* | **14** | Methods for estimating or comparing measures of diagnostic accuracy | 8 |
| | **15** | How indeterminate index test or reference standard results were handled | 7 |
| | **16** | How missing data on the index test and reference standard were handled | 7 |
| | **17** | Any analyses of variability in diagnostic accuracy, distinguishing pre-specified from exploratory | 8 |
| | **18** | Intended sample size and how it was determined | 6 |
| **RESULTS** | | | |
| *Participants* | **19** | Flow of participants, using a diagram | NA |

| | | | |
|---|---|---|---|
| | **20** | Baseline demographic and clinical characteristics of participants | 8 + 26/27 (Table 1) |
| | **21a** | Distribution of severity of disease in those with the target condition | NA |
| | **21b** | Distribution of alternative diagnoses in those without the target condition | NA |
| | **22** | Time interval and any clinical interventions between index test and reference standard | NA |
| *Test results* | **23** | Cross tabulation of the index test results (or their distribution) by the results of the reference standard | 28/29 (Table 2) |
| | **24** | Estimates of diagnostic accuracy and their precision (such as 95% confidence intervals) | 28/29 (Table 2) |
| | **25** | Any adverse events from performing the index test or the reference standard | NA |
| **DISCUSSION** | | | |
| | **26** | Study limitations, including sources of potential bias, statistical uncertainty, and generalisability | 14 |
| | **27** | Implications for practice, including the intended use and clinical role of the index test | 14/15 |
| **OTHER INFORMATION** | | | |
| | **28** | Registration number and name of registry | NA |
| | **29** | Where the full study protocol can be accessed | NA |
| | **30** | Sources of funding and other support; role of funders | NA |

61
